# Hospital mortality in COVID-19 patients in Belgium treated with statins, ACE inhibitors and/or ARBs

**DOI:** 10.1101/2021.03.24.21252687

**Authors:** Geert Byttebier, Luc Belmans, Myriam Alexander, Bo E.H. Saxberg, Bart De Spiegeleer, Anton De Spiegeleer, Nick Devrecker, Jens T. Van Praet, Karolien Vanhove, Reinhilde Reybrouck, Evelien Wynendaele, David S. Fedson

## Abstract

The COVID-19 pandemic has disrupted life throughout the world. Newly developed vaccines promise relief to people who live in high-income countries, although vaccines and expensive new treatments are unlikely to arrive in time to help people who live in low-and middle-income countries. The pathogenesis of COVID-19 is characterized by endothelial dysfunction. Several widely available drugs like statins, ACE inhibitors (ACEIs) and angiotensin receptor blockers (ARBs) have immunometabolic activities that (among other things) maintain or restore endothelial cell function. For this reason, we undertook an observational study in four Belgian hospitals to determine whether in-hospital treatment with these drugs could improve survival in 959 COVID-19 patients. We found that treatment with statins and ACEIs/ARBs reduced 28-day mortality in hospitalized COVID-19 patients. Moreover, combination treatment with these drugs resulted in a 3-fold reduction in the odds of hospital mortality (OR=0.33; 95% CI 0.17-0.69). These findings were in general agreement with other published studies. Additional observational studies and clinical trials are needed to convincingly show that in-hospital treatment with statins, ACEIs/ARBs, and especially their combination saves lives.

## Introduction

The global COVID-19 pandemic has caused massive social, economic and political distress. Many of those infected with the SARS-CoV-2 virus, especially children and younger adults, remain asymptomatic. In contrast, severe COVID-19 largely affects those who are older, especially those with underlying co-morbidities [1].

The global response to the COVID-19 pandemic has been focused on traditional public health measures (e.g., social distancing, wearing masks, etc.) and the rapid development of new vaccines and treatments. Although these efforts promise some relief, COVID-19 will still kill thousands of people worldwide in the months ahead.

The pathogenesis of COVID-19 is characterized (among other things; see below) by endothelial dysfunction [2, 3]. Several inexpensive drugs like statins, angiotensin converting enzyme inhibitors (ACEIs) and angiotensin receptor blockers (ARBs) have immunometabolic activities that maintain or restore endothelial cell function [4]. For this reason, we have undertaken an observational study to determine whether in-hospital treatment with these drugs could improve COVID-19 patient survival.

### The pathogenesis of COVID-19

The pathogenesis of COVID-19 is slowly coming into view and has been extensively reviewed [2, 5-22]. COVID-19 is characterized by severe inflammation, altered interferon, complement and epigenetic responses and dysregulation of innate and adaptive immunity. These changes can lead to acute respiratory distress syndrome (ARDS) and multi-organ failure [5-9], a reflection of renin-angiotensin and endothelial dysfunction that is central to COVID-19’s pathogenesis [2, 3, 10-16]. Endothelial damage is directly responsible for microvascular immunothrombosis and thromboembolism seen in the lungs and other organs of many COVID-19 patients who have died [17-22].

### Treating COVID-19

Many studies of new treatments for severe COVID-19 have focused on new or repurposed antiviral drugs [23-27]. Compared with patients with mild disease, virus loads are higher in those with severe or fatal disease [28, 29]. When clinical deterioration develops, however (usually in the second week of illness), virus loads are generally lower than when symptoms first developed [30, 31].

Most studies of antiviral treatments of hospitalized COVID-19 patients have been disappointing. One of the largest studies was the Solidarity Trial conducted by the World Health Organization (WHO). None of the drugs tested, including remdesivir, improved COVID-19 patient survival [32]. Convalescent plasma [33] and monoclonal antibody preparations [34] have also been tested in hospitalized patients with severe disease. Like antiviral treatments, the trial results have been similarly disappointing. Moreover, the inevitability of SARS-CoV-2 mutations (e.g., 501Y.V2., B.1.1.1.7) jeopardizes the future effectiveness of antibody treatments (and even vaccines) that target the spike protein of the virus [35-37].

Another approach to treating COVID-19 (and other pandemic diseases) focuses on targeting the host response to infection, not the virus [38]. The most prominent drug tested thus far has been dexamethasone, one of several inexpensive corticosteroids that have long been studied for treating patients with sepsis and ARDS. In a randomized controlled trial (RCT), dexamethasone improved survival in COVID-19 patients requiring mechanical ventilation or oxygen treatment but failed to improve survival in those not requiring oxygen treatment [39]. A meta-analysis by a WHO working group concluded that corticosteroid treatment improved survival in COVID-19 patients with severe disease [40]. Nonetheless, questions have been raised about the utility of corticosteroids in these patients [41].

Another approach to treating the host response has used monoclonal antibodies (usually tocilizumab) that antagonize IL-6. Two RCTs have shown that tocilizumab treatment modestly reduced COVID-19 mortality [42, 43] but other studies have shown it did not improve survival [44-46]. Its utility for COVID-19 treatment remains uncertain [47].

Among other drugs considered candidates for host response treatment are statins, ACEIs, and ARBs [4, 48]. Like dexamethasone, these drugs are produced as inexpensive generics, are familiar to practicing physicians and are widely available in resource-poor countries. They have pleiotropic activities on inflammation, affect innate and adaptive immunity, and actively counteract endothelial dysfunction [3, 4, 48-52]. Considered individually, these drugs are safe when given to patients with acute critical illness [53, 54]. Cardiovascular investigators have long known that they are more effective when given in combination than when they are given by themselves [55]. More than a decade ago, statins were proposed to treat pandemic influenza [56] and in 2014 a statin/ARB combination was used to treat Ebola patients in Sierra Leone [4, 57]. In early 2020, combination treatment was proposed for COVID-19 patients [58].

Several studies have described risk factors (e.g., age, co-morbidities) for mortality among patients hospitalized with COVID-19, although most reports provide little detail on the effects of in-hospital treatment [59]. We sought to determine the effectiveness of in-hospital treatment with statins alone, ACEIs/ARBs alone, or a combination of statins and ACEIs/ARBs in reducing 28-day mortality in COVID-19 patients.

## Methods

We undertook a retrospective observational case-control study of the effectiveness of inpatient treatment with statins and/or ACEIs/ARBs on 28-day hospital mortality in 959 COVID-19 patients admitted consecutively to four Belgian hospitals during the first pandemic wave (1 March to 31 July 2020). We used the anonymized and standardized records of the minimal datasets for each hospital. The ethics committees of the four hospitals gave their approvals for the study.

Using propensity scores [60-62], we matched all treated and untreated patients (PSM; 1:1) according to age, sex, hospital size, and underlying co-morbidities (ischemic heart disease, heart failure, stroke, hypertension, chronic kidney disease, diabetes, COPD, asthma, and nicotine use; Table 1). We treated sex as binary variable (M/F) and constrained matching to an exact match between treated and untreated patients. We combined patients treated with ACEIs, ARBs, or both into one group (ACEIs/ARBs) because of small numbers. We evaluated the odds ratios (ORs) for 28-day hospital mortality in patients treated with statins alone, ACEIs/ARBs alone and combination treatment with statins and ACEIs/ARBs using conditional logistic regression with conditioning on matched sets. The use of PSM allowed us to minimize the potential effects of confounding variables and balance patient characteristics in treated and control groups. The Logit Propensity Score summarized the collective influence of all covariates on treatment assignment for each treated group (statins alone, ACEIs/ARBs alone and statins + ACEIs/ARBs combinations) and their corresponding untreated control groups. The Standardized Mean Differences (SMDs) between patients in treated and untreated control groups were also calculated.

**Table 1.** Patient baseline characteristics

| Variable | All patients<br>(n=959) | No statin<br>and no<br>ACEI/ARB<br>(n=571) | Statin<br>only<br>(n=178) | ACEI/ARB<br>only<br>(n=91) | Statin +<br>ACEI/ARB<br>(n=119) |
| --- | --- | --- | --- | --- | --- |
| Age (years):<br>mean/median<br>(minimum-<br>maximum) | 69.2/73.0<br>(0-98) | 64.9/69.0<br>(0-98) | 76.2/77.0<br>(40 - 96) | 73.2/75.0<br>(44-94) | 76.2/75.0<br>(41-97) |
| <40 years of age | 65<br>(6.8%) | 65<br>(11.4%) | 0<br>(0%) | 0<br>(0%) | 0<br>(0%) |
| 40-60 years of age | 187<br>(19.5%) | 156<br>(27.3%) | 11<br>(6.2%) | 16<br>(17.6%) | 4<br>(3.4%) |
| ≥60 years of age | 707<br>(73.7%) | 350<br>(61.3%) | 167<br>(93.8%) | 75<br>(82.4%) | 115<br>(96.6%) |
| Male | 523<br>(54.5%) | 303<br>(53.1%) | 104<br>(58.4%) | 41<br>(45.05%) | 75<br>(63.0%) |
| Hypertension | 329<br>(34.3%) | 127<br>(22.2%) | 62<br>(34.8%) | 58<br>(63.7%) | 82<br>(68.9%) |
| Asthma | 33<br>(3.4%) | 21<br>(3.7%) | 5<br>(2.8%) | 4<br>(4.4%) | 3<br>(2.5%) |
| COPD | 161<br>(16.8%) | 82<br>(14.4%) | 38<br>(21.3%) | 17<br>(18.7%) | 24<br>(20.2%) |
| CKD | 216<br>(22.5%) | 100<br>(17.5%) | 51<br>(28.6%) | 25<br>(27.5%) | 40<br>(33.6%) |
| Diabetes | 188<br>(19.6%) | 61<br>(10.7%) | 52<br>(29.2%) | 20<br>(22.0%) | 55<br>(46.2%) |
| Nicotine | 45<br>(4.7%) | 23<br>(4.0%) | 11<br>(6.2%) | 5<br>(5.5%) | 6<br>(5.0%) |
| History of nicotine | 143<br>(14.9%) | 63<br>(11.0%) | 37<br>(20.8%) | 15<br>(16.5%) | 28<br>(23.5%) |
| IHD | 95<br>(9.9%) | 30<br>(6.2%) | 36<br>(20.2%) | 6<br>(6.6%) | 23<br>(19.3%) |
| Stroke/TIA | 42<br>(4.4%) | 19<br>(3.3%) | 9<br>(6.1%) | 2<br>(2.2%) | 12<br>(10.0%) |
| HF | 115<br>(12.0%) | 49<br>(8.6%) | 27<br>(15.2%) | 9<br>(9.9%) | 30<br>(25.2%) |

ACEI = angiotensin converting enzyme inhibitor; ARB = angiotensin receptor blocker; COPD = chronic obstructive pulmonary disease; CKD = chronic kidney disease; IHD = ischemic heart disease; TIA = transient ischemic attack; HF = heart failure

To account for competing risks (i.e., the chance of hospital discharge competing with the risk of death), we also calculated cumulative incidence functions (CIFs) and the associated Gray’s tests to compare treated and untreated control groups [60-62].

## Results

Among all 959 patients (mean age 69.2 years, 54.5% men) (Table 1), 388 (40.6%) were treated as inpatients with one or more of these drugs (statins and/or ACEIs/ARBs). In the treated group, 49.4% had underlying cardiovascular diseases or hypertension. Among all patients, 707 were ≥ 60 years of age (Table 1). In this older age group, 357 (50.5%) were treated with these one or more of drugs, accounting for 92.0% of all 388 patients who received these treatments.

Among all patients, 150 (15.6%) died in hospital within 28 days (Table 2). All but three deaths occurred among patients ≥ 60 years of age. Unadjusted mortality rates in this age group were lower in patients who had been treated with one or more of these drugs compared with those not treated.

**Table 2.**
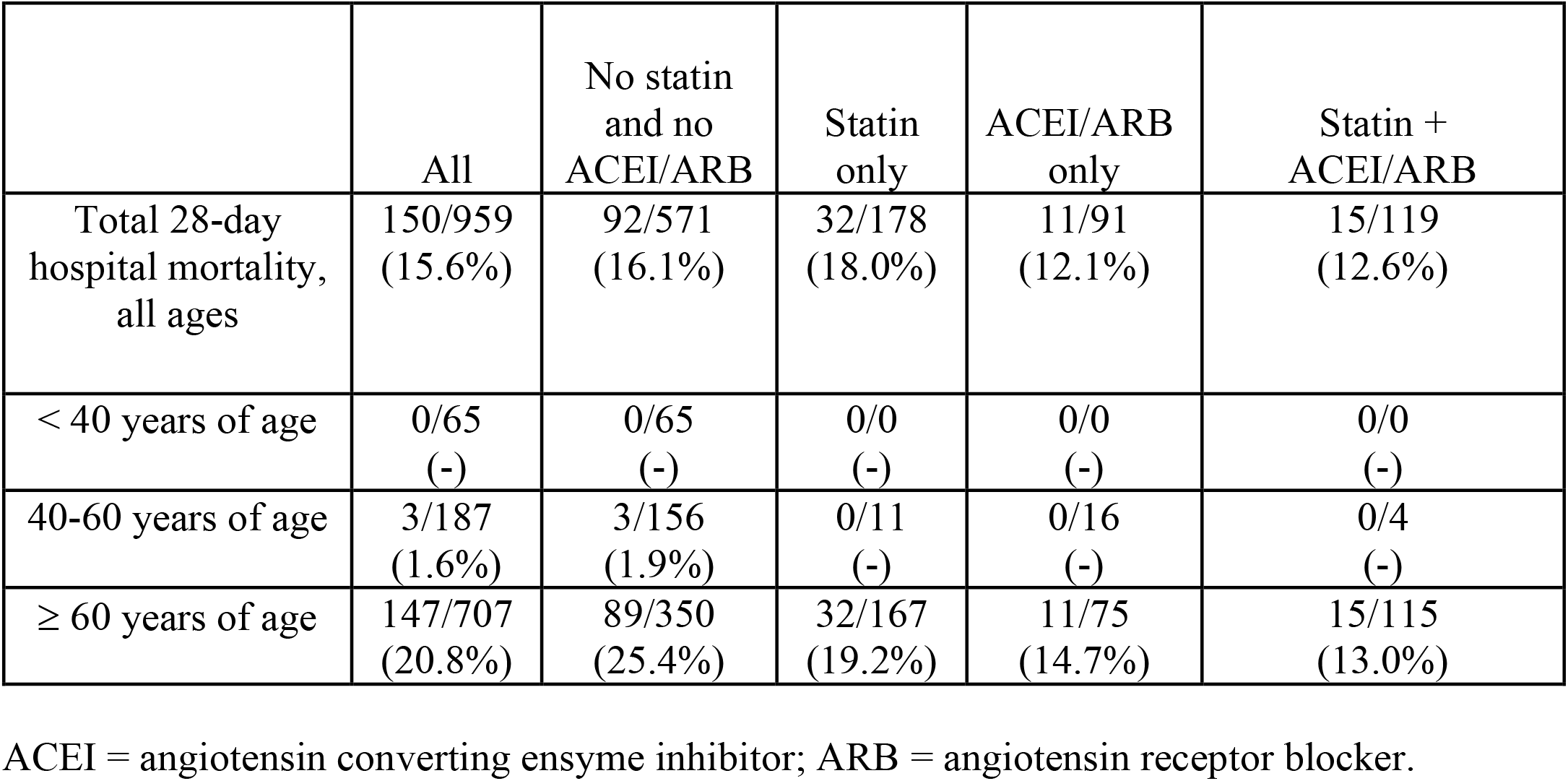
Unadjusted in-hospital 28-day mortality by age and treatment group.

Figure 1 illustrates the Standardized Mean Differences (SMDs) after the PSM procedure. The logit propensity score (“Logit Prop Score”) summarizes the collective influence of all observed covariates on treatment assignment. In addition to the SMDs for all observations and the matched observations, Figure 1 also shows the SMDs for “region observations”. This is a selection of observations whose propensity scores (or equivalently, logits of propensity scores) lie in the region of common support for the treated and control groups. This region is the largest interval that contains propensity scores (or logits of propensity scores) for subjects in both groups.

**Figure 1.**
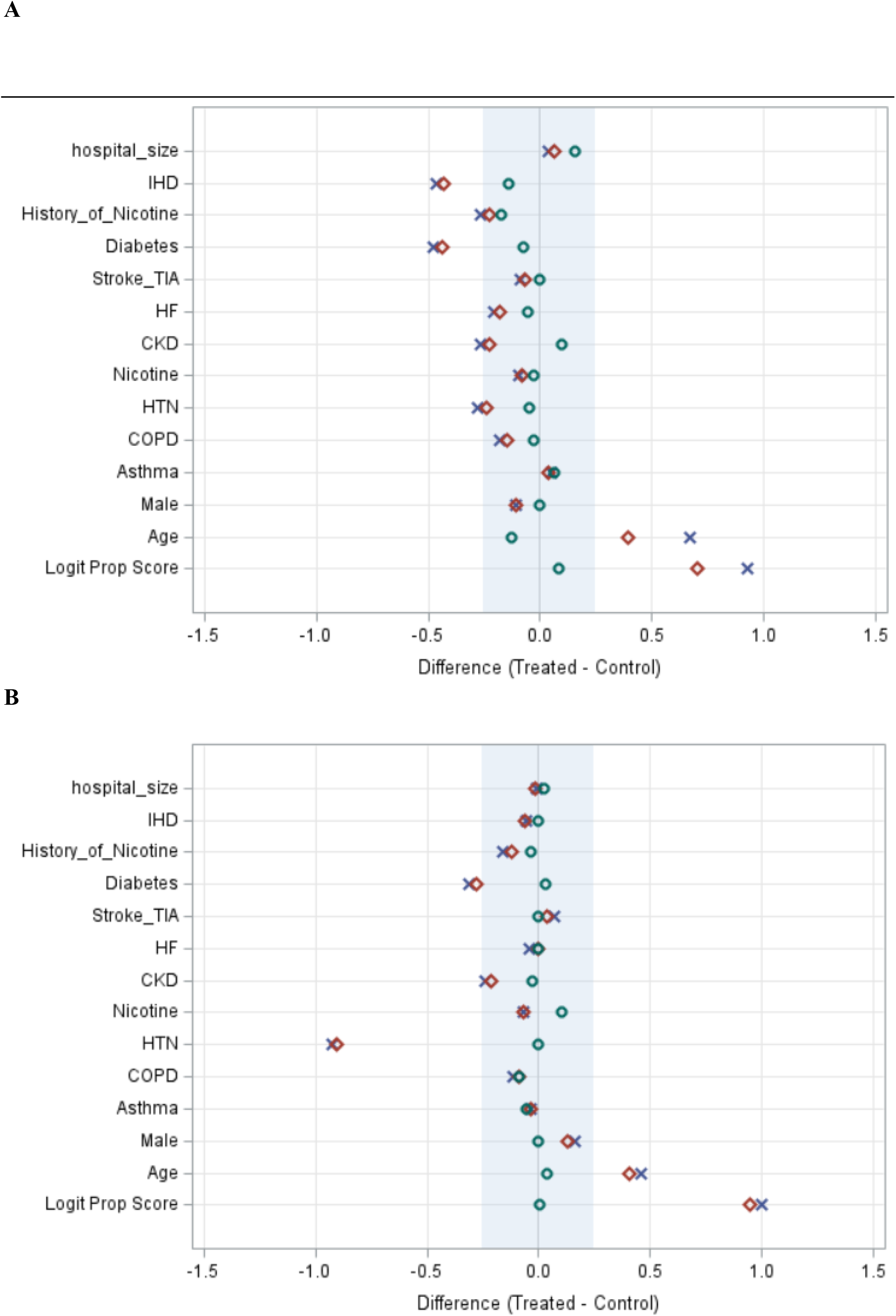
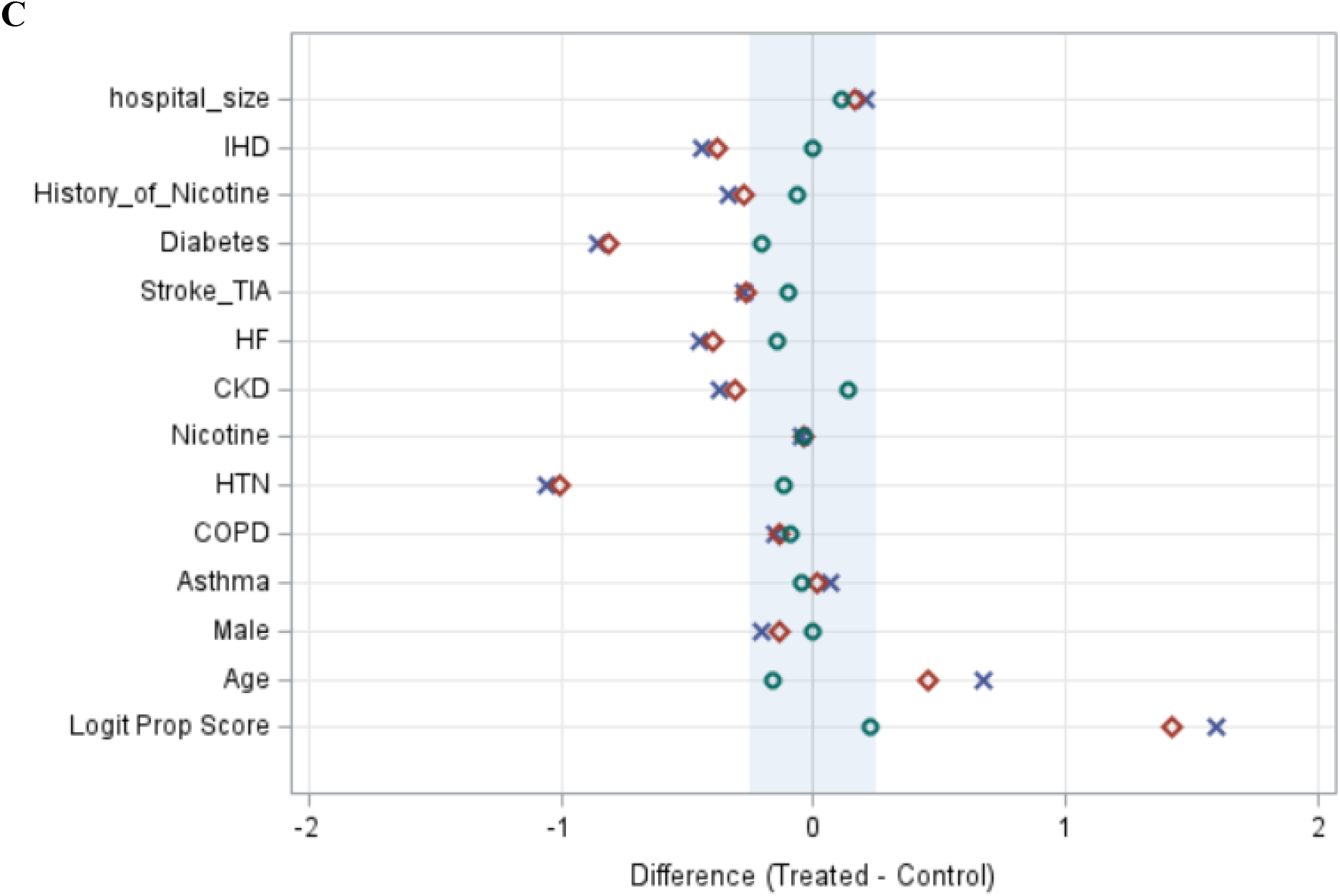
Standardized Mean Differences (SMDs) before and after propensity score matching (PSM) for treatment with statins alone (A); ACEIs/ARBs alone (B); and combination treatment with statins + ACEIs/ARBs (C) versus the control group (no treatment with statins or ACEIs/ARBs) for each treated group. The shaded areas indicate SMDs <0.25. SMDs were < 0.10 for 9/13 variables for statins alone (A), 12/13 for ACEI/ARB alone (b) and 6/13 for combination treatment (C). x = all observations *before* matching, ◊ = region observations (see text for definition), o = propensity score observations *after* matching. See text for definition of Logit Prop Score. Abbreviations for co-morbidities are defined in the legend for Table 1.

Using propensity score matching (PSM) in all matched sets, the SMDs between treated and untreated control groups were considered acceptable if <0.25, although many individual SMDs were <0.10 (Figure 1). For combination treatment with statins + ACEIs/ARBs (117 matched sets), the adjusted (conditional logistic regression) odds ratio (OR) for 28-day hospital mortality was 0.33 (95% CI 0.17-0.69; p=0.002, SMD = 0.22) (Table 3). For treatment with statins alone (177 matched sets) and ACEIs/ARBs alone (90 matched sets), the adjusted ORs were 0.56 (95% CI 0.39-0.93, p=0.024, SMD = 0.08) and 0.52 (95% CI 0.23-1.17, p=0.11, SMD = 0.006), respectively (Table 3).

**Table 3.** Observational studies of 28-day hospital mortality following in-hospital treatment of COVID-19

| Study/type of study (ref) | In-hospital treatment | Adjusted HR/OR | 95% CI | p value |
| --- | --- | --- | --- | --- |
| BHS/<br>PMS-CCS (a) | statins | 0.56 | 0.39-0.93 | 0.02 |
|  | ACEIs/ARBs | 0.52 | 0.23-1.17 | 0.11 |
|  | statins + ACEIs/ARBs | 0.33 | 0.17-0.69 | 0.002 |
| Zhang/<br>PSM-CCS (64) | statins | 0.58 | 0.43-0.80 | 0.001 |
|  | statins + ACEIs/ARBs | 0.32<sup>b</sup> | 0.12-0.82 | 0.018 |
| Mallow/<br>cohort (65) | statins | 0.54 | 0.49-0.60 | 0.001 |
| Fan/<br>PSM-CCS (66) | statins | 0.25 | 0.07-0.92 | 0.037 |
| Masana/<br>GM-CCS (67) | statins | 0.60 | 0.39-0.92 | 0.02 |

Abbreviations: BHS = Belgian Hospital Study; HR = hazard ratio; OR = odds ratio; CI = confidence interval; PSM-CCS = propensity score matched case-control study; ACEI = angiotensin converting enzyme inhibitor; ARB = angiotensin receptor blocker; GM = genetics matched.
<sup>a</sup> This study.
<sup>b</sup> Patients with statins + ACEIs/ARBs treatment were compared with those with statins + no ACEIs/ARBs treatment.

Using Gray’s test, the CIF, which accounted for the difference in mortality between the statins + ACEIs/ARBs vs no statins and no ACEIs/ARBs, was highly significant; p=0.0040 (Fig. 2C). The CIFs for mortality did not reach significance for statins alone vs no statins (p= 0.0670; Fig. 2A) and for ACEIs/ARBs alone vs no ACEIs/ARBs (p=0.1928; Fig. 2B).

**Figure 2.**
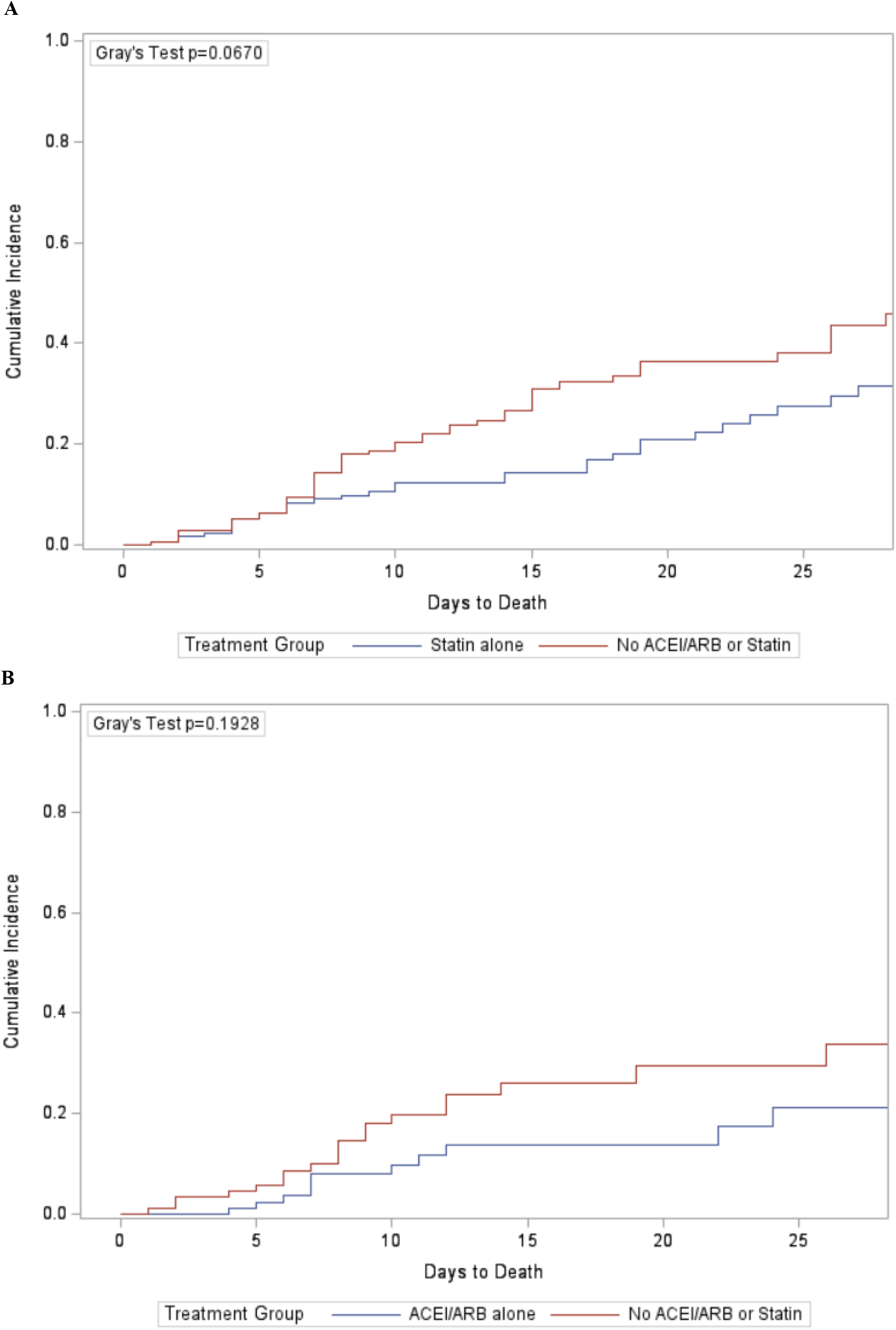
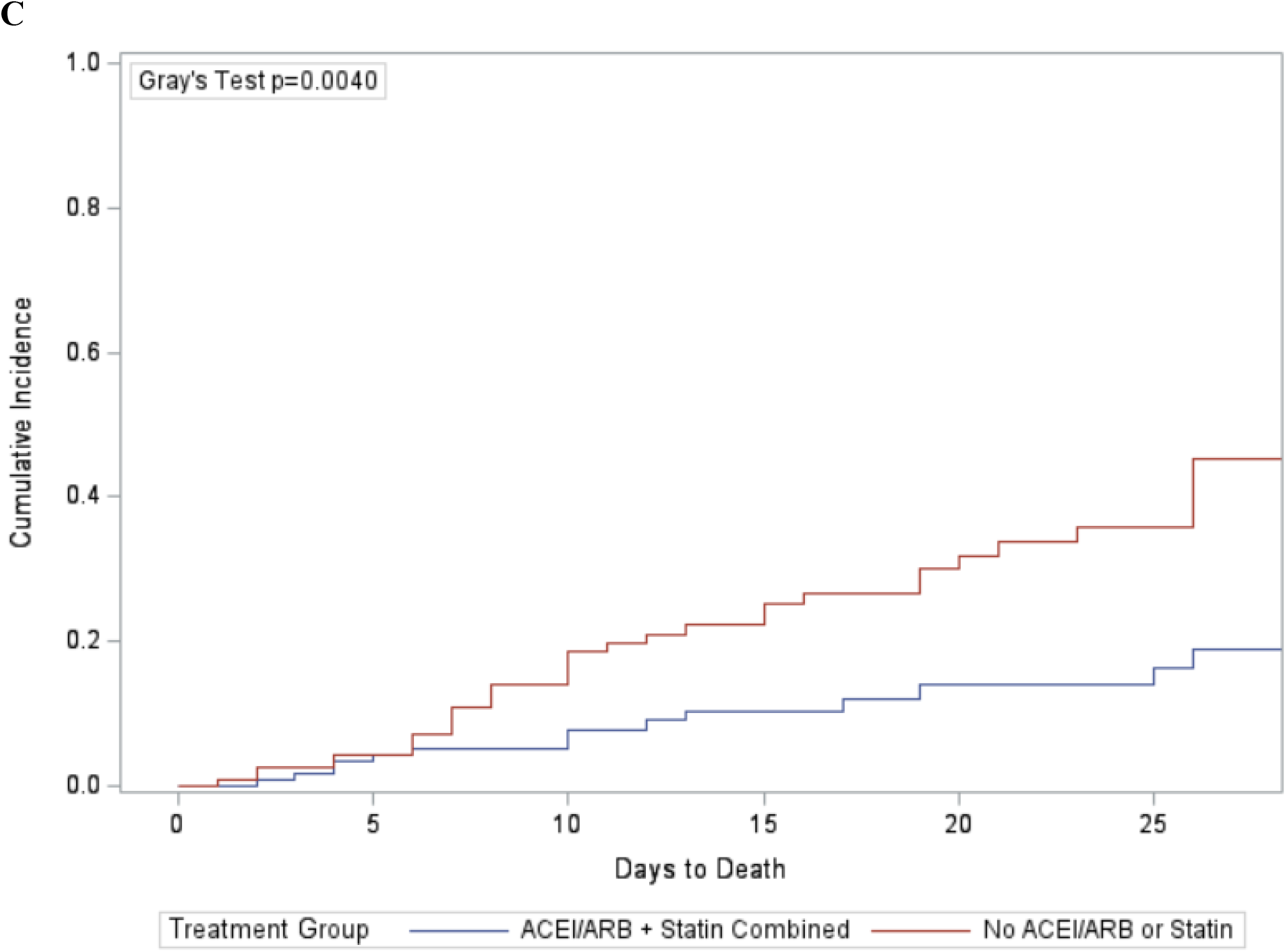
Cumulative Incidence Functions (CIFs) for days to death after PSM for treatment with statins alone (A), ACEIs/ARBs alone (B) and combination treatment with statins + ACEIs/ARBs (C). Because the number of PSM-matched patients in each treated group was different, the corresponding number of patients in each PSM-matched untreated control group was different.

In all treatment groups, the beneficial effects of treatment were evident beginning 7 to 8 days after hospital admission (Figure 2).

## Discussion

Newly developed vaccines are currently being used in a limited number of high-income countries. These countries account for most of the advanced purchase orders for COVID-19 vaccines [63]. However, in low- and middle-income countries people will have little access to COVID-19 vaccines until well after most of them have been infected. Similarly, most of the studies of new treatments involve patented agents that will be expensive and in short supply. These agents are unlikely to be widely available in resource-poor countries.

The results of our study compare favorably with those of other observational studies of inpatient treatment of COVID-19 patients with statins and ACEIs/ARBs (Table 3) [64-68]. Similar to our study, these reports show that in-hospital treatment with statins alone or ACEIs/ARBs alone improved survival. In addition, we observed that combination treatment with statins and ACEIs/ARBs was associated with a 3-fold reduction in the odds of 28-day hospital mortality (OR =0.33); Table 3).

We did not have information on whether our treated COVID-19 inpatients had also been treated as outpatients, although it is likely that many who had been treated as outpatients continued their treatment after hospital admission. However, several individual studies [e.g., 68-72] and meta-analyses [73, 74] of statin treatment have yielded conflicting results. In these studies, treatment was ascertained largely on the basis of outpatient (not inpatient) information. For example, in a study of 247 statin-treated COVID-19 patients, 46% of treatments were initiated after hospital admission and yet 29% of all statin treatments were discontinued because of elevated liver function or creatine kinase tests [66]. Another study showed that outpatient statin treatment was associated with decreased COVID-19 mortality, although only 77% of outpatient-treated patients continued statin treatment after hospital admission [71]. If statins are withdrawn at the time of hospital admission, their beneficial effects are likely to be reduced [75, 76]. Thus, statin treatment should be continued (or started) after hospital admission [76, 77]. Whether the same is true for ACEI and ARB treatment is uncertain [78, 79]. Clinical guidelines now recommend that outpatient statin and ACEI/ARB treatments should be continued in hospitalized COVID-19 patients [80, 81].

The endothelial dysfunction that characterizes COVID-19 is associated with microvascular immunothrombosis and venous thrombosis, which are the result of widespread disturbances in coagulation [16-21]. Many patients with severe COVID-19 also have a variety of functional autoantibodies that cause thrombosis [82, 83] and compromise interferon activity [84]. Drugs such as statins, ACEIs and ARBs were initially developed to treat cardiovascular diseases. Because of their pleiotropic activities [49], some investigators believe they could be repurposed to treat the many immunometabolic derangements that characterize COVID-19 [4, 50-52]. For example, randomized controlled trials have been initiated in ICU-admitted and non-ICU-admitted COVID-19 patients to evaluate whether therapeutic anticoagulation is more beneficial than prophylactic anticoagulation. Recently, enrollment of ICU (but not non-ICU) patients in these studies was “paused” for reasons of futility and safety [85]. It is not yet known which of these patients might have been receiving other treatments. Statins have demonstrated anticoagulant effects [86, 87] and counteract the effects of autoantibodies on endothelial cells [88]. There is little information on the anticoagulant effects of ACEIs and ARBs, although ARBs have beneficial effects on many of the signaling pathways involved in coagulation (see Table 2 in reference 4).

Treatment with statins, ACEIs/ARBs, or a combination of both might be considered for persons who test positive for COVID-19 but have yet to develop symptomatic disease or be admitted to hospital. Preventing the development of symptomatic disease might even prevent “long COVID” [89]. In a small study of 154 nursing home residents in Belgium who were screened and found to be COVID-19-positive, those who were being treated with statins had significantly fewer episodes of symptomatic COVID-19 [90]. Although residents taking ACEIs and ARBs tended to have fewer episodes of symptomatic disease, the results were not statistically significant. Similarly, those taking statins, ACEIs and ARBs had fewer episodes of serious COVID-19 requiring hospitalization, but again the results failed to reach statistical significance.

Most investigators who have reported observational studies showing that statin and/or ACEI/ARB treatment reduces COVID-19 mortality have called for randomized controlled trials (RCTs) to confirm their findings [e.g., 64, 66, 67, 70, 71]. Some investigators insist that RCTs are essential [91, 92]. Most of these drugs are widely available, inexpensive generics that have been repurposed for COVID-19 treatment. Unfortunately, only 7% of the studies registered on Clinicaltrials.gov are focused on cardiovascular treatments and most of them are single center studies of ACEIs and ARBs [93]. Recently, a proposal for an RCT that included combination treatment (NCTTO4343001) was withdrawn because funding could not be obtained. To our knowledge, no RCT of combination treatment of COVID-19 patients is currently underway.

Our observational study has several limitations. The sample size (959 patients) was small; our findings will need to be replicated in larger studies. We could not evaluate separately the effects of ACEI and ARB treatment, although investigators have reported greater effectiveness for ACEI compared with ARB treatment [94]. We did not correct for immortal time bias. We did not have access to information on outpatient treatments and thus could not evaluate whether outpatient statin treatment was continued in the hospital or initiated only after hospital admission. We could not evaluate deaths that occurred after 28 days or that might have occurred after hospital discharge. We also did not evaluate the effects of treatment on length of stay, ICU admissions or mechanical ventilation.

We are currently expanding our study to include the second pandemic wave (1 August to 31 December 2020) and additional outcomes (e.g., ICU admissions, mechanical ventilation). We will also evaluate separately the contributions of ACEI and ARB treatments alone or in combination with statins to reducing in-hospital COVID-19 mortality and their effectiveness in treating asymptomatic (but PCR test-positive) patients.

Observational studies and RCTs both have their places in the hierarchy of research designs [95]. However, if there are no RCTs [96], physicians and health officials will be forced to consider the trade-off between pandemic learning and doing [97]. In this circumstance, they might have to rely on well-conducted observational studies alone to determine whether combination treatment of the host response in COVID-19 patients is effective [98].

## Conclusion

Our observational study in Belgium has shown that COVID-19 patients treated in-hospital with statins in combination with ACEIs/ARBs experienced a 3-fold reduction in the odds of 28-day hospital mortality. Given the absence of RCTs of combination statin + ACEI and statin + ARB treatment, investigators should undertake additional observational studies (as well as RCTs) of these treatments in hospitalized COVID-19 patients. In light of the recent increase in COVID-19 cases and hospitalizations and the strong signal of potential effectiveness shown in our study, they should be able to do so quickly. Moreover, regardless of underlying co-morbidities and unless contraindicated, physicians might also consider combination treatment with these drugs for all hospitalized COVID-19 patients.

## Data Availability

Data referred to in this article has been anonymized and is kept with the study investigator.

## Acknowledgements

The authors thank clinicians in the four Belgian hospitals for their support: AZ-Delta (Roeselare), AZ-Vesalius (Tongeren), AZ-Sint-Jan (Brugge) and RZ-Heilig Hart (Tienen).

